# Co-Immune: a case study on open innovation for vaccination hesitancy and access

**DOI:** 10.1101/2021.03.29.20248781

**Authors:** Camille M. Masselot, Bastian Greshake Tzovaras, Chris L.B. Graham, Gary Finnegan, Rathin Jeyaram, Isabelle Vitali, Thomas E. Landrain, Marc Santolini

## Abstract

**Background:** The rise of major complex public health problems, such as vaccination hesitancy and access to vaccination, requires innovative, open and transdisciplinary approaches. In spite of this, institutional silos, paywalls and lack of participation of non-academic citizens in the design of solutions hamper efforts to meet these challenges. Against this background, new solutions have been explored, with participatory research, citizen science, hackathon and challenge-based approaches being applied in the context of public health.

**Objectives:** Our ambition was to develop a framework for creating citizen science and open innovation projects that address the contemporary challenges of vaccination in France and around the globe.

**Methods:** We designed and implemented Co-Immune, a programme created to tackle the question of vaccination hesitancy and access to vaccination through an online and offline challenge-based open innovation approach. The programme was run on the open science platform Just One Giant Lab.

**Results:** Over a 6-month period, the Co-Immune programme gathered 234 participants of diverse backgrounds and 13 partners from the public and private sectors and organized 8 events to facilitate the creation of 20 new projects as well as the continuation of 2 existing projects to address the issues of vaccination hesitancy and access, ranging from app development and data mining to analysis and game design. In an open framework, the projects made their data, code, and solutions publicly available.

**Conclusion:** Co-Immune highlights how open innovation approaches and online platforms can help to gather and coordinate non-institutional communities in a rapid, distributed and global way towards solving public health issues. Through the ideas of hackathons and other contest approaches, such initiatives can lead to the production and transfer of knowledge, creating novel solutions in the public health sector. The example of Co-Immune contributes to paving the way for organisations and individuals to collaboratively tackle future global challenges.

## Introduction

### Background

As the world faces a rise in the number of complex challenges that threaten the resilience of our economic, environmental and social systems, we observe a shift towards more collaboration and openness in the way science and innovation is performed [1–3], bringing closer governments, civil society, and the private sector. Examples of this include the efforts made to accelerate society’s progress towards Sustainable Development Goals (SDG) [4] and the fight against pandemics such as COVID-19 [5]. Yet, access to vaccines and vaccination hesitancy remains one of the complex challenges to be addressed to achieve universal health coverage [6].

In the last decade, the World Health Organization (WHO) Global Vaccine Action Plan 2011-2020 [7] committed 140 countries and 290 organizations to promoting and prioritising greater collaboration between governments, NGOs, the private sector and all citizens to address outbreaks of vaccine-preventable diseases. Indeed, immunization is one of the most cost-effective interventions to protect oneself and others from infectious diseases [7] saving between two million and three million lives per year [8]. Yet, the annual death toll for vaccine-preventable diseases stands at 1.5 million [7] whereas the WHO listed vaccine hesitancy among the Top 10 Global Health Threats for 2019 [9]. Continuing global efforts to leave no one behind may be a long-standing challenge [10] when new information technologies and social media platforms are both part of the problem [11], and solution.

In response, a number of new digital and open innovation initiatives have been launched: the WHO has developed the Vaccine Safety Net [12], a network of websites about vaccination; health authorities in Canada have developed a schools-based quiz to educate children about immunology and vaccines [13]; Finland is testing a computer game to communicate the benefits of HPV vaccination [14]; a project in India uses digital necklace to record children’s immunization history [15], and a global Vaccination Acceptance Research Network has been established [16].

The number, relevancy, sustainability and impact of these types of initiatives could be amplified by fostering increased collaboration with non-academic citizens in the creation and development of solutions in an open innovation framework [17]. This is the gap that Just One Giant Lab (JOGL) is proposing to fill with the Co-Immune programme.

In response to such complex, multi-scale problems, the practice of science is changing. Research demonstrates that intensity and diversity of collaboration positively affect the quality [18] and productivity [19] of research while positively impacting the knowledge integration from participants [20]. Likewise, participant transdisciplinarity [21] seems critical to generating innovative outcomes [22] and dealing with complex real world problems [23]. Such mechanisms are often at play in the field of citizen science, promising to transform the knowledge generation landscape by tapping into networks of non-academic citizens [24,25] in a new social contract for this kind of research [26]. Citizen science has the potential to expand the number of individuals contributing knowledge and ideas, transform how hypotheses are generated and datasets are analysed. Such approaches have already been applied to investigate individual diseases through patient-led research [27,28] and public health challenges such as the epidemiology of cancer [29–31].

Other approaches to create and develop knowledge and solutions to complex challenges are slowly entering the mainstream. In particular, hackathons, challenge-based approaches, and the participation of citizens in science have been flourishing over the last two decades [32], especially within the natural sciences [33] and more recently, medical sciences, public health and population-health research [34,35].

Hackathons are short, intensive, and collaborative events that are designed to prototype solutions addressing a specific problem. They originated in the early 2000s in digital and tech fields and have been adapted to address more complex challenges in global health [36–38]. Such initiatives are not without pitfalls: they suffer, by design, from the lack of paths to sustainability for the projects they launch [39]. In response to such criticisms, there are increasing efforts, such as “Make the Breast Pump not Suck” hackathon and “Trans*H4CK”, to improve hackathon methodology by working directly with affected communities [40]. Several initiatives such as MIT collaborative design studio provide insights on hackathon methods [41] to facilitate better hackathons [42,43]. More recently, multiple entities have engaged in organizing hackathons to address the COVID-19 crisis [44,45].

Challenge-based approaches, providing frameworks for learning while solving real-world issues, have also been on the rise in global health and proven to be efficient to generate innovative solutions and incentivize mass community engagement [46]. For example, the potential of participative models to address complex questions, and the power of contests to offer a structure that catalyses this work, has been exhibited by the “Epidemium” initiative on cancer epidemiology [47].

Despite the numerous tools and technologies created to facilitate collaboration in citizen science projects, challenges remain. These include the issue of the complementarity, coherence and diffusion of these initiatives [32] to efficiently address international policies and local needs, as often the local adoption of hackathon solutions remains low [39].

The promotion of transdisciplinarity and citizen science in an open innovation framework, coupled with methods such as hackathons and a challenge-based approach therefore represent an opportunity to address current complex challenges of vaccination, that would overcome the limits of either solution alone. In this article, we describe the design, implementation, and outputs of “Co-Immune”, a collaborative open innovation programme run in 2019 to address vaccination hesitancy and access to vaccination.

### Objectives

Co-Immune’s ambition was to develop a framework for creating and developing citizen science and open innovation projects addressing the contemporary challenges of vaccination in France and around the globe. The programme had four specific objectives: (a) to foster a collaborative, open and transdisciplinary dynamic (b) to promote the emergence of accessible knowledge and innovative solutions (c) to support participants in the elaboration and development of their project and (d) to disseminate the outputs and results in an open science framework.

## Methods

### Design

The overall programme duration was 10 months (March 2019 to January 2020), divided into 6 months of preparation and 4 months of roll-out of activities that included offline and online events, support for the development of citizen science projects, and assessment and awards for projects participating in the challenge-based competition. The main output of the programme were projects, categorized as leading to a) knowledge production, if they performed data analysis or generated new knowledge whether it is specific to context or generic [48]; and/or b) knowledge “transfer” [49], and/or solutions such as hardware, software, and interventions.

Co-Immune was coordinated online through the platform of JOGL (app.jogl.io) and supported by 13 partners from the public and private sector (Supplementary Table 1).

The governance of Co-Immune was designed to provide freedom for projects to develop innovative solutions while ensuring their compliance with local and international regulations and consideration of ethical and scientific integrity. To this end, we constituted an independent Committee for Ethics, Science and Impact (CESI), which issued an opinion on the rules of participation in the programme and validated the strategic orientation of the programme. Public health priorities were identified based on a literature review and divided between two main challenges to streamline participants’ work: vaccination coverage, and vaccination hesitancy. They were then validated by the CESI. In addition, through a series of semi-structured interviews, experts at the 7th Fondation Merieux Vaccine Acceptance conference [50] identified eight specific issues to address and potential room for solutions. The CESI also participated in the co-elaboration of the assessment grid, used as a base to grant non-monetary prizes to projects in December 2019.

### Participant recruitment

Participants were recruited through our network of partners from around the globe and social media communication. Participation was open to everyone above the age of 18, if they agreed to follow the participation rules validated by the CESI. Participants could take the role of “project leaders” and “contributors’’.

### Just One Giant Lab (JOGL) platform

Co-Immune participants used the JOGL platform to document their projects and recruit collaborators throughout the course of the programme. JOGL is a decentralized mobilization platform designed for use in collaborative research and innovation (Figure 1a). Within JOGL platform users can create a profile and declare their skills. Once registered, they can create or join projects, follow the activity of other members, post on their project feed and comment on other posts. They can also highlight needs for a project they are part of, specifying skills that can help to solve them. We compared the JOGL features to other online platforms for citizen science, social networking and science/publishing through a cluster analysis (Figure 1b & Supplementary information), indicating it is functionally similar to other platforms in the space and is suitable to hold a citizen science programme such as Co-Immune.

**Figure 1:**
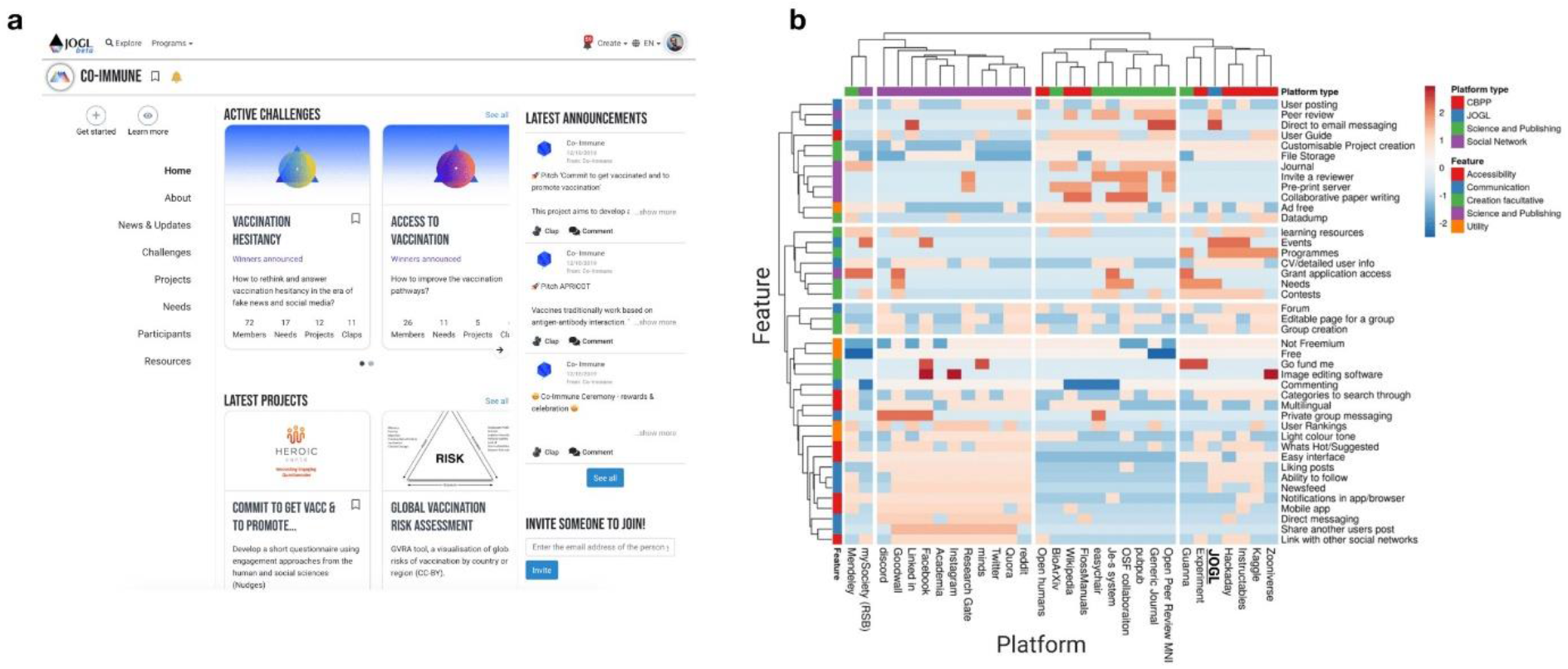
Overview of the JOGL platform. **a** screenshot of the JOGL platform (app.jogl.io). **b** Heatmap of feature presence across popular online tools. For each platform (columns), we numerically encode the presence (1) or absence (0) of each feature (rows). We then compute for each element a Z-score by standardizing values across platforms, represented here by the color spectrum: blue-low to red-high. CBPP-Citizen based peer production network/citizen science platforms.

### Implementation

The Co-Immune programme was realised through an interrelated and interacting set of technological and social features (see Figure 2). Our coordination team implemented the larger framework (events, online platform, contest approach) and helped to recruit a community of partners and participants which interacted with each other and were supported in their efforts through the high-level design features. With support of the governance structure of the Co-Immune programme the individual projects managed to provide outputs that included knowledge production and transfer, and solutions such as hardware, software, and interventions.

**Figure 2:**
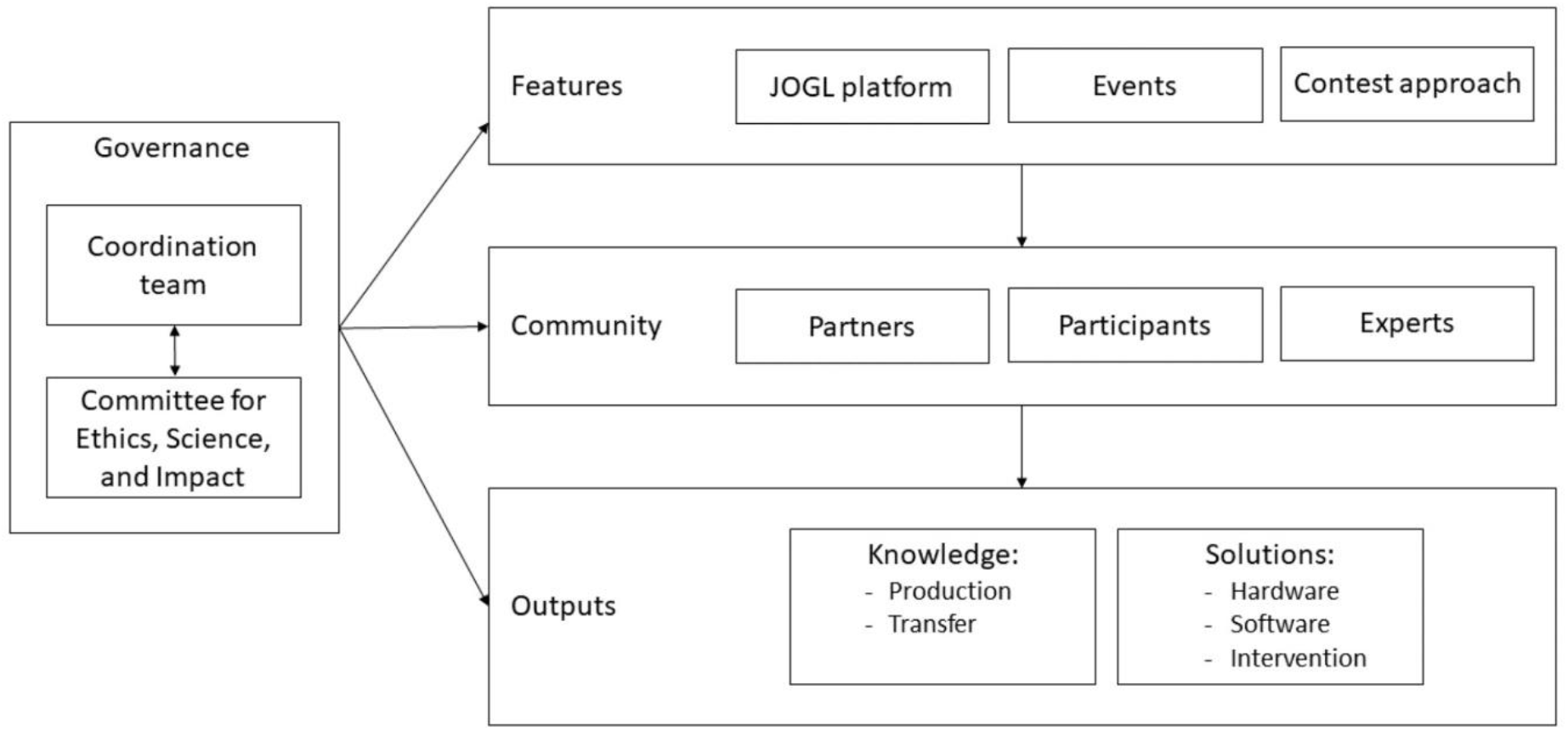
Workflow of the Co-Immune programme design.

### Building an open community

To build the community, we contacted organisations involved in a wide range of domains before the launch of the programme, creating a first pool of contributing professionals and students. We also recruited participants via the organization of events, typically in the evening, aimed at creating projects, fostering collaboration among participants to address project needs, and provide mentorship. To facilitate the coordination of the community, all participants were required to use the JOGL platform to describe their projects, form teams, list their needs and initiate collaboration.

We organized 8 offline and online events between October and December 2019 (Table 1). Participants for events were recruited through social media and mailing lists leveraging our network of partners. Among the four onsite events that were organized, two were hackathons aimed at motivating participants to join the programme, while the other two were aimed at fostering collaboration around the most advanced projects. Their median duration was 3 hours.

**Table 1:** Co-Immune events

| Name | Mode<br>Type<br>Location | Duration<br>(hours) | Objective | Design<br>Supporting partners | Number of<br>Participant<br>s |
| --- | --- | --- | --- | --- | --- |
| Launch | Offline<br>Ceremony<br>CRI, Paris | 3 | Gather the initial community | Presentation of the programme design, features, timeline and partners, networking. | 60 |
| OpenJOGL<br>"Co-Immune" | Online | 1 | Q&A session on the programme | Presentation of Co-Immune, questions and answers. | 3 |
| Sprint<br>"Open Data" | Offline<br>Hackathon<br>CRI, Paris | 2.5 | Build community, create projects, create data repositories | Statement of the problem (videos of experts), team formation and effort, mentoring, publication of results on the JOGL platform.<br>Supported by: CRI, CorrelAid, | 25 |
| OpenJOG<br>"Vaccination awareness escape game" | Online | 1 | Foster collaboration around single project | Pitch of the project and its needs, feedback from experts, questions and answers. | 7 |
| Sprint<br>"Project creation" | Offline<br>Hackathon<br>CRI, Paris | 4 | Build community, create multidisc. projects | Statement of the problem (videos of experts), ice breaker and multidisciplinary team formation and effort, mentoring, presentation of results and vote for the most promising projects, publication of results on the JOGL platform, networking.<br>Supported by: CRI, Epitech, Wild code school, CorrelAid, Excelya | 22 |
| Sprint<br>"Open Data" | Offline<br>Hackathon<br>Wild code school, Paris | 3 | Accelerate the development of projects related to data science | Selection of a project by participants among the 2 choices available, team formation and effort, mentoring, presentation of results and publication on the JOGL platform, networking.<br>Supported by: Wild code school, CorrelAid, Excelya | 15 |
| Sprint<br>"Open Data" | Offline<br>Hackathon<br>Epitech, Paris | 3 | Build the community, create projects, accelerate the development of one project using twitter data | Statement of the problem, selection of a project by participants among the 4 choices available including one already existing project, team formation and effort, mentoring, presentation of results, vote for the most promising project, publication of results on the JOGL platform, networking.<br>Supported by: Epitech, Kapcode, Excelya, Correlaid | 35 |
| OpenJOGL<br>"HERA - a<br>mobile health<br>platform to<br>improve<br>refugees'<br>health " | Online | 1 | Foster<br>collaboration<br>around single<br>project | Pitch of the project and its needs,<br>feedback from experts, questions and<br>answers. | 7 |
| OpenJOGL<br>"Better<br>documentatio<br>n for better<br>collaboration" | Online | 1 | Help teams<br>documenting<br>their project in<br>the most open<br>and<br>reproducible<br>way | Expert presentation on best practices for<br>documenting open science projects,<br>presentation of Co-Immune expectations<br>for documentation, questions and<br>answers. | 13 |
| Closing<br>ceremony | Offline<br>Ceremony<br>CRI, Paris | 2 | Close the Co-Immune<br>programme | Presentation of the main outputs of the<br>programme, reward of the best projects. | 70 |

The facilitation of the hackathon-style events relied on the use of participatory and collective intelligence design and problem-solving techniques [51]. In particular, participants were encouraged to form multidisciplinary teams including both professionals and students.

Partners co-organized offline events and mobilized experts of their network to support the participants. Three partners in Paris – Epitech, the Wild code school and the Center for Research and Interdisciplinarity (CRI) – co-organized and hosted events for their students respectively in their engineering, coding, and life science and education schools. Other partners – such as Kapcode, Excelya, and CorrelAid – mobilized their teams to act as mentors during these events. A total of 14 mentors attended events, five came to more than one event.

In addition, we organised four one-hour online events. The first was an opportunity to share information about Co-Immune with people around the globe. Another event discussed best practices to document open-science projects. Finally, two events focused on the resolution of needs of single projects (Table 1).

### Co-Immune challenge-based approach and project assessment

The challenge-based nature of the programme was designed to be an incentive for teams and participants to continue developing their projects after hackathon events or create their project on JOGL at any other time. To be eligible for a prize, a project was required to have created a comprehensive description of their initiative on the JOGL platform and a video pitch. This material was provided to experts in charge of the assessment. We considered “expert” all the members of the CESI and experienced professionals of a certain field who attended events and provided technical guidance to teams as “mentors”.

The assessment was also designed to be an opportunity for learning and growth. In addition to grades, teams received detailed feedback on their project. Project assessment was performed through a grid co-developed by JOGL and the CESI.

The assessment grid was based on a literature review of project evaluation standards and consisted of 10 questions graded from 0 to 5 (Supplementary Table 2). Three areas were assessed: the approach, the implementation strategy, and the impact. First, the assessment of the approach included: i) Clarity and relevance of the problem and alignment with the programme scope ii) Fit between approach/methodology and problem statement iii) Innovation potential: the project introduces ground-breaking objectives, novel concepts or approaches. Second, the implementation strategy was assessed following the criteria: i) State of progress towards set goal (state of advancement); ii) Clarity and relevance of the timeline and needs for future (major tasks, milestones); iii) Project actively engages and aligns with all relevant stakeholders. Finally, the assessment of the impact covered: i) Clarity and relevance of the criteria used to measure impact; ii) To what extent does the project considers its ecosystem (ecological, environmental, ethical and social considerations); iii) Sustainability and scalability of the project in the long term; iv) Open and reproducible dissemination strategy. For each of these 3 categories, JOGL awarded a prize to the project with the best score based on the grades given by reviewers. Additionally, a Grand Prize was given to the project with the overall highest score. JOGL provided visibility while two partners also provided rewards to a project of their choosing.

### JOGL platform data collection and analysis

Participants gave their professional background, their skills and employment status on JOGL. This data was used to evaluate the composition of the community. All users who joined JOGL during the span of the programme were considered to be participant of Co-Immune, as it was the only ongoing programme and all outreach activities were related to it.

To better understand how skills were related across participants, we used a network approach to assess similarity between skills and get further insights about the global diversity of the community. In this network approach, each declared skill is a node and the considered skills as linked if they co-occur in a participant. Links are then weighted by the number of participants they co-occur in. Gephi 0.9.2 was used to represent the network in Figure 5, and its modularity algorithm was used with default parameters to compute communities.

## Results

### Community growth through events

During the programme, 234 participants signed up to the platform (Figure 3a). The participant growth was mostly linear over the lifespan of the programme (2019-07-10 to 2019-12-18), suggestive of a potential for continued growth if the programme had continued. The growth rate outside of events, at around 1 per day (between 0.86 - 0.98 users/day), is consistent with the pre-kickoff growth rate (0.94 users/day). This highlights the importance of events (dashed lines in Figure 3a) for driving participant enrolment, with the 4 offline events accounting for 45% of the growth. In total, offline events were responsible for the generation of 82% (18/22) of projects. The rest consists of four projects created on the platform outside of events and two already existing prior to the programme.

**Figure 3:**
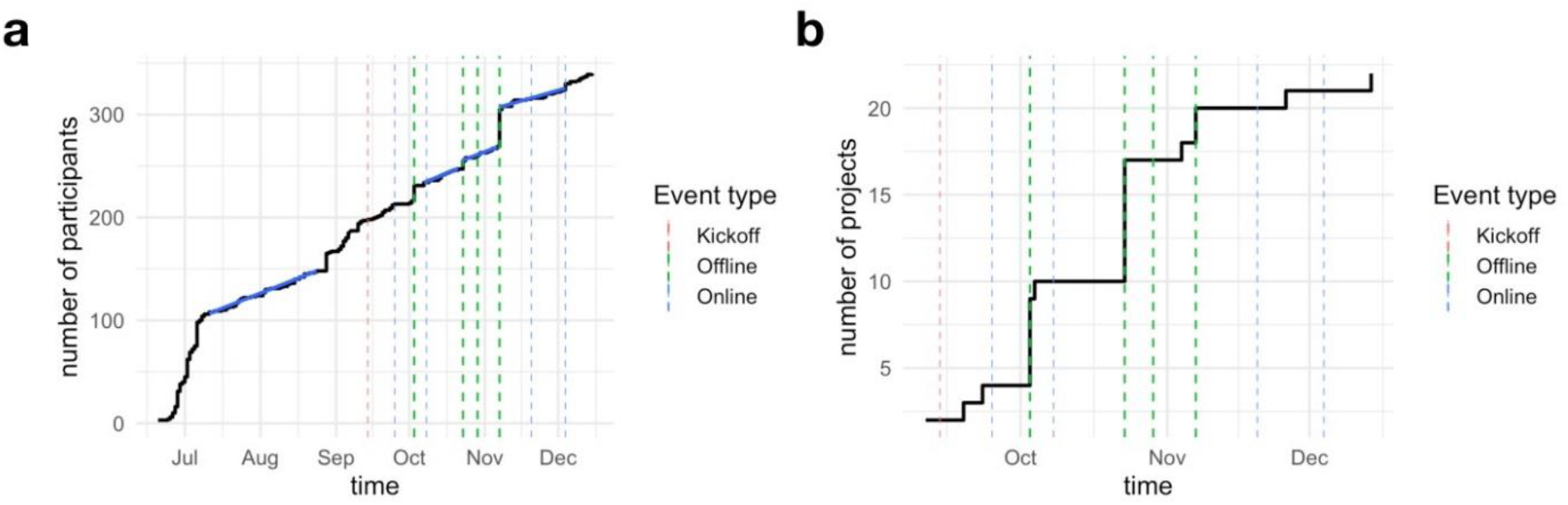
Growth of the **a** number of participants and **b** number of projects over the lifespan of the programme. Dashed bars show when events for community facilitation where held (green: offline events, blue: online events, red: kickoff meeting). Blue lines give a linear fit during the corresponding periods, showing stable growth pre- and post-kickoff.

### Participant skills and backgrounds: a transdisciplinary community

Out of the 234 participants, 187 declared their job category. The community was composed of a mix of students (67/187, 36%) and workers (94/187, 50%), most of whom were full time (86%) (Figure 4a). Other categories included ‘between jobs’ (1), ‘non-profit’ (12) and ‘for profit’ (3). While 60% of the participants were based in Paris, approximately 40% of all enrolled participants came from regions including the rest of Europe, the Americas, Africa & Asia.

**Figure 4:**
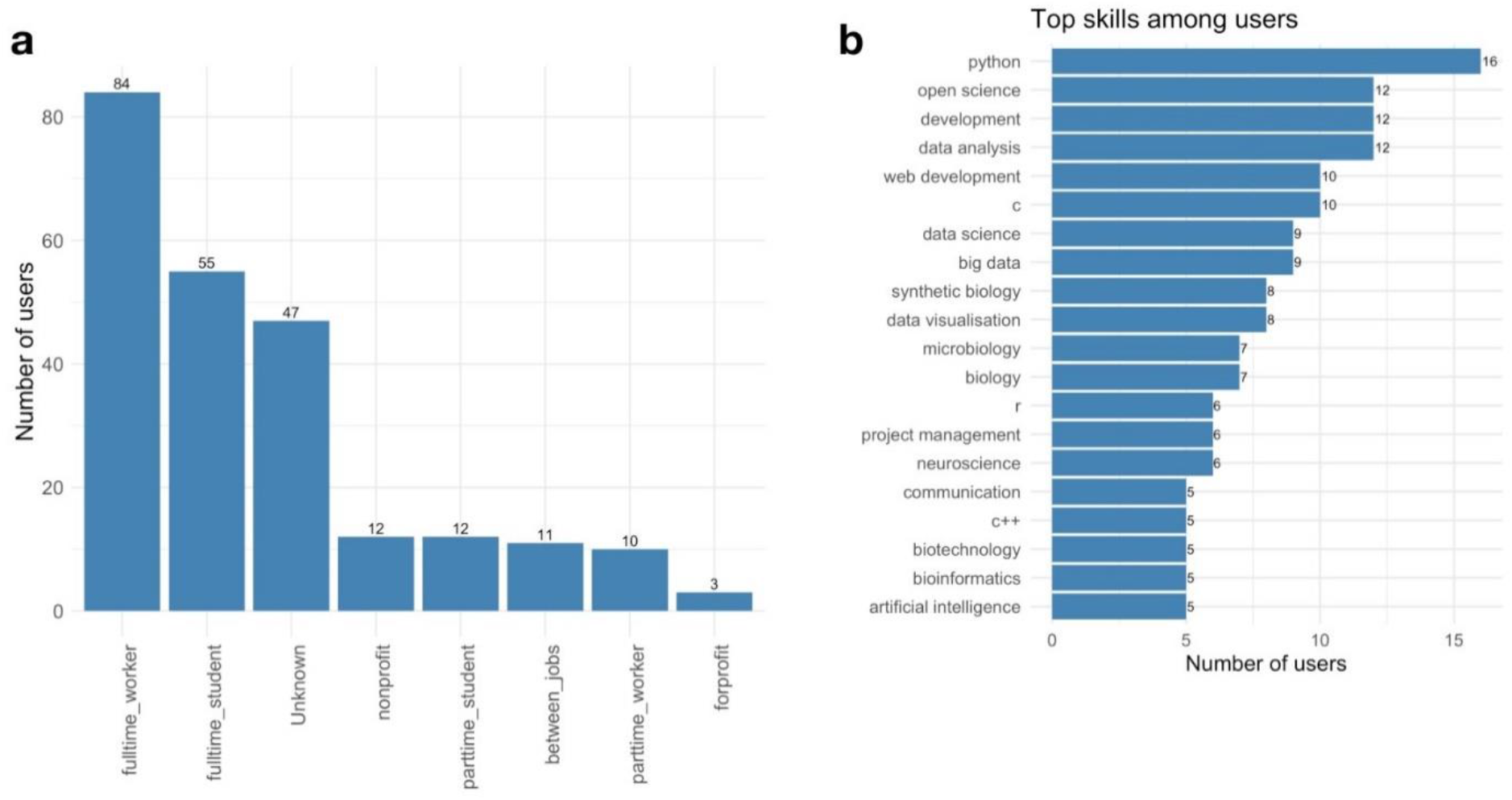
An overview over the Co-Immune community. **A** Participants category. **B** The 20 most represented skills in the Co-Immune community.

The 234 participants specified a total of 492 unique skills (median 3 skills per participants). We observe a high representation of data science and coding alongside biology, which altogether relate to the technical skills emphasized during the programme (Figure 4b.). The skill network shows that the community spans a vast interdisciplinary landscape: from open science to open data and coding, and from project management to biology. The network exhibits a giant component of 416 interconnected skills (84% of all skills) (Figure 5). This giant component was in turn analysed for topological modules using modularity maximisation (see Methods). Modularity maximization resulted in the identification of 12 modules corresponding to sets of skills that tend to co-occur more than with other skills. Since these skills are linked through the participants who share them, they can be understood as “participant types’’ constitutive of the Co-Immune community.

**Figure 5:**
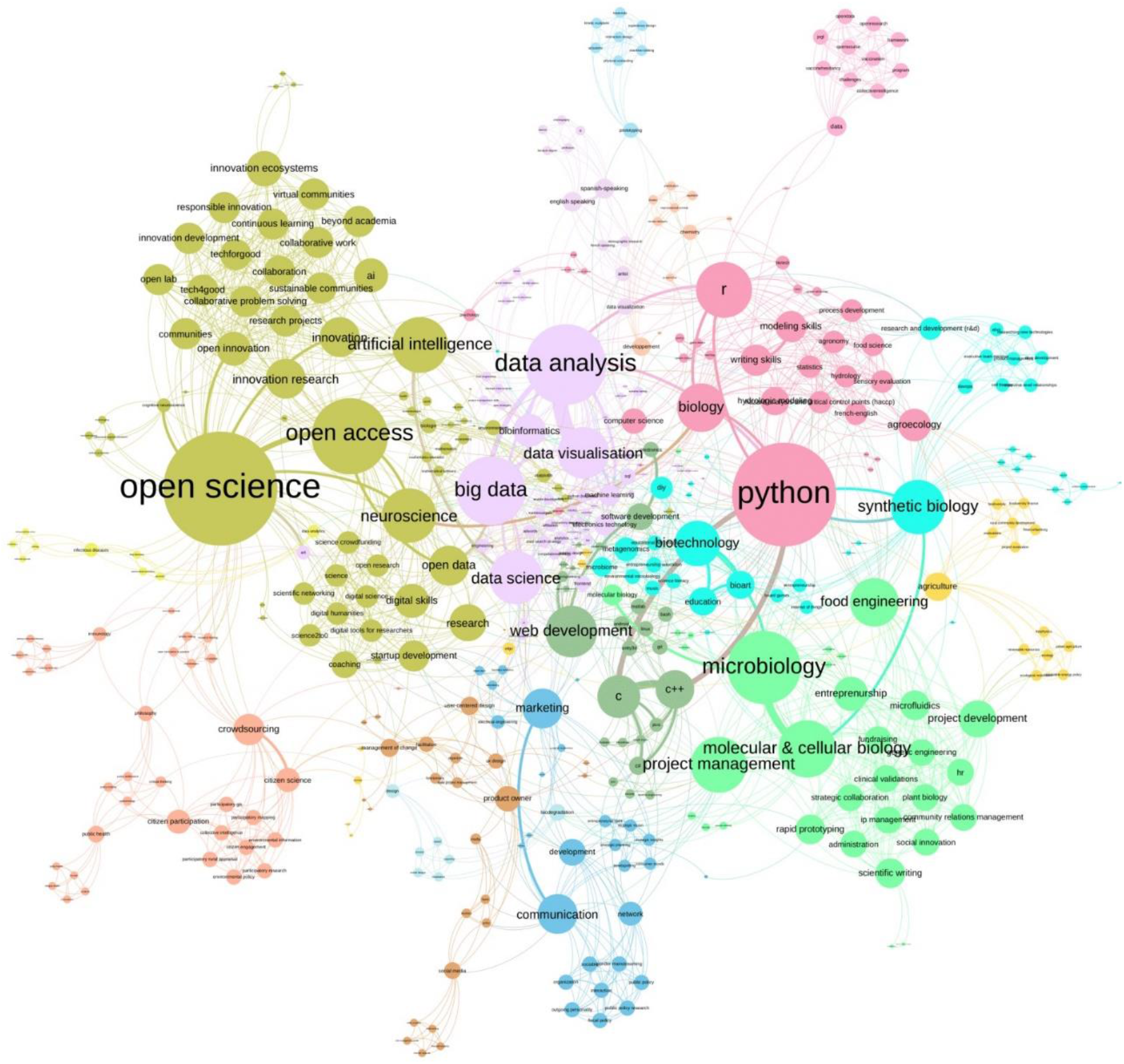
Skill map of the Co-Immune community. Skills are linked if they appear in the profile of the same participant. Link weight indicates the number of participants sharing the skills. Node size indicates weighted degree.

### Co-Immune partners & experts

The 13 partners operated in the health, technology, and social sectors, and included research, innovation, and education organizations, as well as professional networks, incubators, and communication specialists (see Figure 6). The number of partners grew over the lifespan of the initiative and were often suggested by existing partners or through connections made during events.

**Figure 6:**
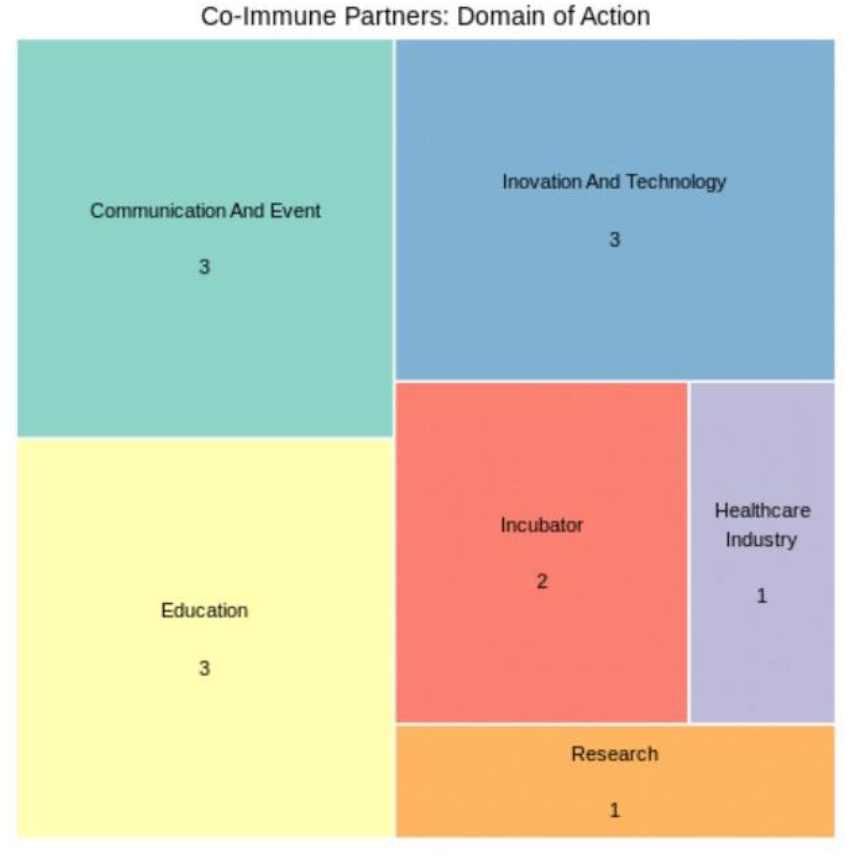
Treemap representing the domain of action of the 13 Co-Immune partners.

In addition to the partners, we mobilized experts including members of the CESI, mentors during events, and interviewees at a conference regarding the challenges to be addressed in the programme. Overall, their domain of expertise ranged from biology, to social sciences, design, technology and data science (Figure 7). A third of them were working as health or public health professionals.

**Figure 7:**
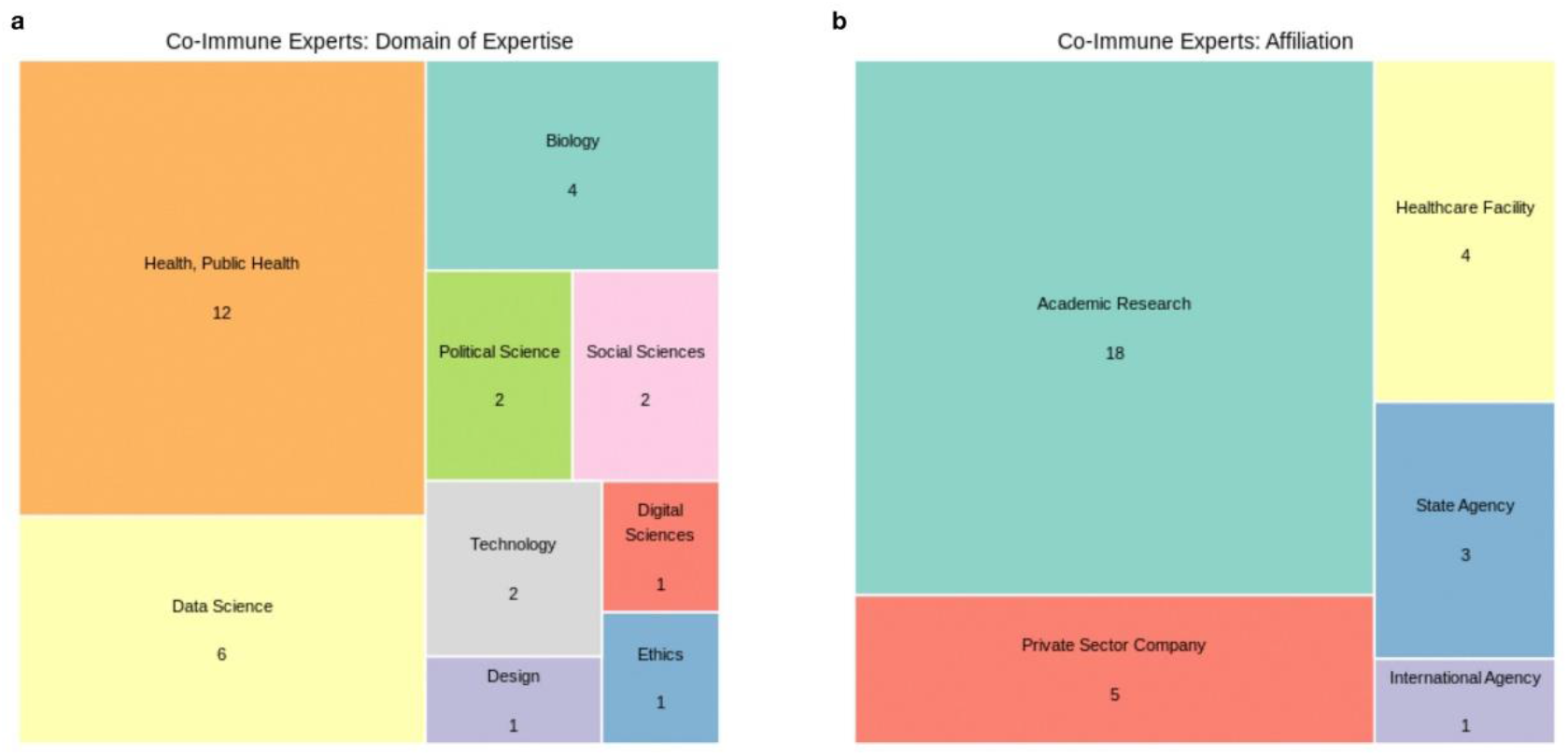
Treemap of the 31 Co-Immune experts: **a** Domain of expertise. **b** Affiliation.

The CESI counted 8 volunteer members and included virologists, pharmacists, health economists, experts in the digital and ethics field and biologists, working at international, national and local levels of the health system. All of them worked for public or nonprofit organisations.

Interviewees were mostly researchers in social sciences and medical practitioners.

### Co-Immune projects & evaluation

A total of 22 projects were created by 20 project leads, with teams of up to 11 members (Table 2). Among these, 15 projects proposed to develop software covering web technologies, mobile apps, algorithms, data lakes, data modeling, analysis and visualisation tools. The 7 other projects included hardware development and interventions involving biotechnologies, game design, behavioral sciences, education and communication. Overall, a third of the projects focused on knowledge transfer.

**Table 2:** Co-Immune project description

| Project name | Project status | Solution category | Summary description |
| --- | --- | --- | --- |
| <u>HERA: a mobile health platform to improve refugees' health</u> | Reviewed<br>Awarded<br>Grand Prize<br>Best approach prize<br>Best impact strategy prize | Software<br>Knowledge transfer | Mobile health app designed for improving the monitoring of vaccination and perinatal health of Syrian refugees in Turkey. It provides recall of vaccines, storage of health data, health promotion (educational content), financial incentives for immunization. |
| <u>Qualitative analysis of Tweets on Vaccination</u> | Reviewed<br>Awarded<br>Partner prize | Software<br>Knowledge production | Web based platform providing real time visualisation and analysis of tweets related to vaccination and vaccination hesitancy. Data analysis included sentiment analysis and network analysis. An area of development was the development of predictive models of epidemic occurrence based on twitter data. |
| <u>HEROIC Santé - Commit to get vaccinated and to promote vaccination</u> | Reviewed<br>Awarded<br>Best implementation strategy prize | Intervention<br>Knowledge transfer | A short questionnaire (7 minutes) using engagement approaches from the human and social sciences, such as "the importance of the source," "voluntary consent," or "fear and danger management," to engage healthcare professionals and users not only to be vaccinated against the flu, but also to promote flu vaccination. |
| <u>Project APRICOT</u> | Reviewed<br>Awarded<br>Partner prize | Hardware | Development of a synthetic biology-based methodology which addresses the evasion mechanisms adopted by the mycobacterium tuberculosis and induce the acceleration of lysosomal biogenesis improve antigen presentation. |
| <u>Vaccination Awareness Escape Game</u> | Reviewed | Intervention<br>Knowledge transfer | Escape Game to raise vaccination awareness among the general population. |
| <u>Harmonize Vaccination</u> | Reviewed | Software<br>Knowledge production | Tool for parsing various formats of vaccination coverage data sets and to visualize them on a common platform. |
| <u>Pass it on: A game about vaccine hesitancy</u> | Reviewed | Software<br>Knowledge transfer | Role-play video game aiming to improve the capacity of health professionals to respond to their patient's hesitation to be vaccinated. |
| <u>Global Vaccination Risk Assessment</u> | Reviewed | Software<br>Knowledge production | A tool to create an overview of risk factors of "not getting vaccinated" by country while looking at the more comprehensive picture. The methodology of this project is based on Fuzzy Logic, multi-criterion analysis, and risk triangle. |
| <u>Immuno</u> | Not reviewed | Hardware<br>Knowledge transfer | Board game providing access to the general public's understanding of medical sciences related to immunisation |
| Vaccine DataDump | Not reviewed | Software<br>Knowledge production | Vaccination related data repository and analysis tool for quick analysis of vaccine related issues. |
| Measuring vaccination hesitancy from social media | Not reviewed | Software<br>Knowledge production | Data analysis of social media (Twitter) to examine if negative sentiment related to vaccination precedes declaration of symptoms and study the relation between vaccination hesitancy and epidemiological outbreaks. |
| Mortality according to access to vaccines | Not reviewed | Software<br>Knowledge production | Data analysis exploring the link between immunisation coverage, mortality rate and distance from health centers. |
| The health system matrices | Not reviewed | Software<br>Knowledge production | Exploratory analysis of the various parameters influencing vaccination coverage over time. |
| Meta immune - Data exploration of existing DB | Not reviewed | Software<br>Knowledge production | Data lake on immunization data. |
| Biloba (projet tiré de) | Not reviewed | Intervention | Intervention incentivising people to increase vaccine uptake through vouchers, supporting the existing mobile App Biloba. |
| Wakuchin Senshi | Not reviewed | Intervention<br>Knowledge transfer | An interactive role play board game to increase the awareness about vaccination among general population |
| Neutralizing information about vaccines | Not reviewed | Software<br>Knowledge transfer | Algorithm parsing web pages, identifying misinformation and trustful content to help users in their health decisions related to vaccines. This also aims to be used by search engines in their recommender systems. |
| Go Viral ! | Not reviewed | Intervention<br>Knowledge transfer | Communication campaign on social media using gamification methods to illustrate contagion among users and thereby increase the awareness of the importance of vaccines. |
| Make Vaccines Affordable | Not reviewed | Software<br>Knowledge transfer | A web based portal with data related to population demand for care in order to negotiate prices of vaccines with suppliers. |
| Analyse de tweets liés à la vaccination | Not reviewed | Software<br>Knowledge production | An analysis of discussion in vaccination related posts on Twitter and their evolution over time. |
| Detect vaccine administration in social media patient data | Not reviewed | Software<br>Knowledge production | A classifier able to detect vaccine administration in tweets related to vaccination |
| Detect vaccine hesitancy in social media patient data | Not reviewed | Software<br>Knowledge production | A classifier able to detect vaccine hesitancy in tweets related to vaccination |

Among the 15-project relying on software technology, 11 projects aimed at contributing to the production of knowledge by facilitating the analysis of publicly available data via the use of parsing tools and the creation of repositories (N=2), the analysis of open data (N=3), the development of machine learning tools to extract and analyse Twitter data related to vaccination hesitancy (N=2), and the production of data visualizations (N=3). In particular, more than 40 datasets were identified and collected by 4 projects created during the data-centered events. In addition, a database of 2,464 tweets (in French) posted over a period of 7 years was made available by a partner and another dataset of 89,979 tweets was gathered by the project ‘Live Twitter Data analysis and visualization on Vaccination’.

Four projects used software for knowledge transfer: For instance, the HERA project provided educational content and health data storage through their mobile app to improve the monitoring of vaccination and perinatal health among Syrian refugees in Turkey. The “Pass it on” project focused on role-playing video games directed to health professionals as another method of knowledge transfer. The “Neutralizing information about vaccines” project implemented an algorithm for parsing web pages, helping citizens identify trustworthy content related to vaccines.

A total of 5 projects focused on different interventions, including raising awareness about vaccination through an escape game (Table 2, “Vaccination Awareness escape Game”); communication campaigns on social media (Table 2, “Go viral!”). The “Heroic Santé” project developed and tested a short questionnaire using engagement approaches from social sciences to engage healthcare professionals and users around the question of flu vaccination (Table 2, “Heroic Santé”). Finally, one team proposed to apply synthetic-biology methods to tuberculosis vaccines (Table 2, “project APRICOT”).

Seven projects provided sufficient documentation on JOGL to be reviewed by the pool of independent experts. In total, 27 reviews were performed, yielding scores ranging from 18 to 32.8 out of a possible total of 45 across the different dimensions that were assessed (approach, implementation strategy, and impact). The average score was 25.1 (+/-6.4).

“HERA: a mobile health platform to improve refugees’ health” was awarded with prizes for best approach (mean score of 11.4/15) and impact (mean score of 14.6/15). “HEROIC Santé - Commit to get vaccinated and to promote vaccination” was awarded the best implementation strategy prize (mean score of 10.33/15).

The projects were globally more successful in terms of approach with an average score of 9.37 out of 15 points (+/-1.79 Stdev). Four projects (Figure 8) had a score higher than 4/5 for the clarity, relevance, and alignment of their problem statement with the programme objectives. For 6 projects the fit between the methods and the projects’ objectives was scored highly by reviewers, with a score of at least 3/5.

**Figure 8:**
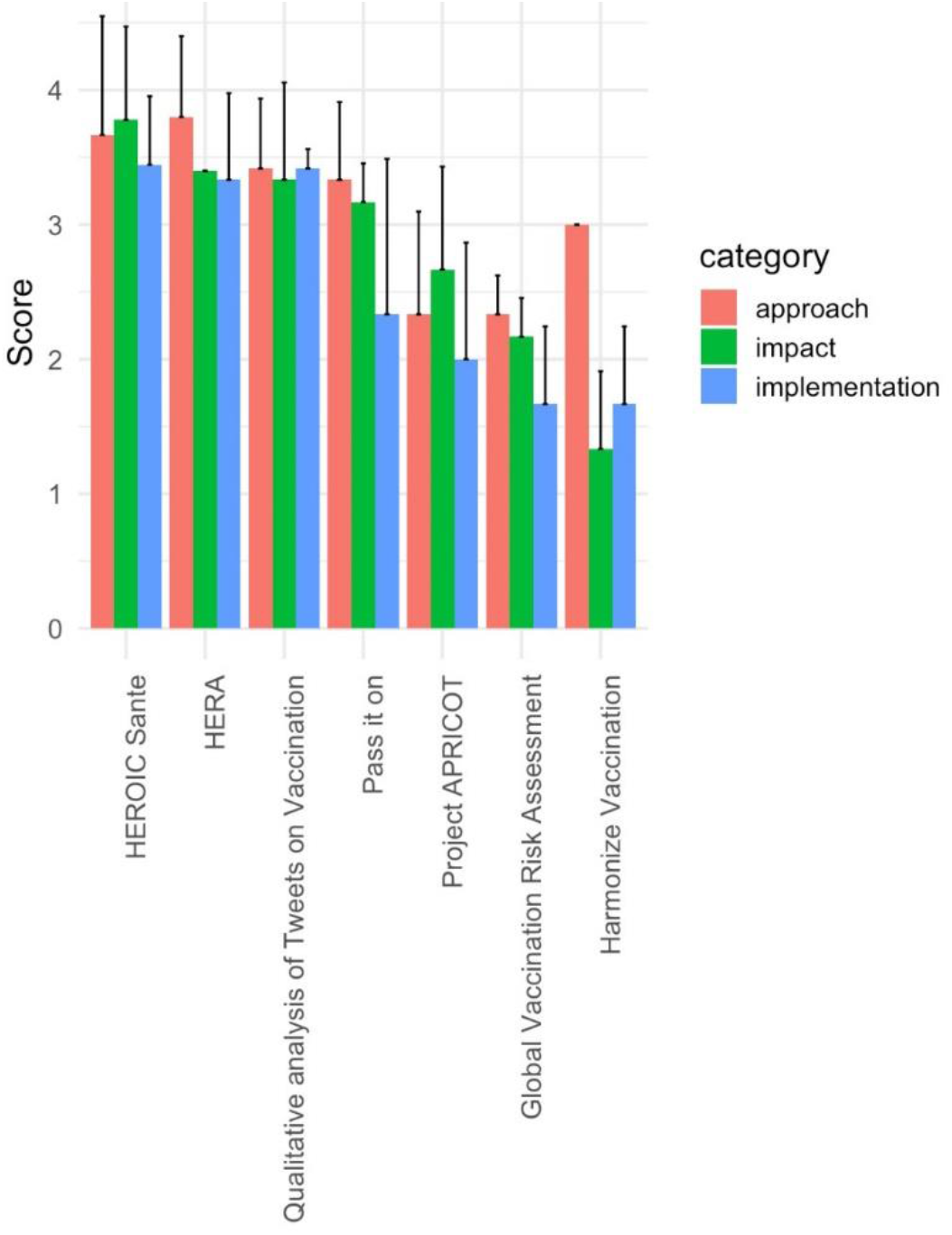
Barplot of review scores per category for all reviewed projects. Bars show average value for all questions related to each category, and error bars represent standard deviation. Projects are shown by decreasing global score.

The implementation strategy score of projects was overall low given the early stage of the projects at reviewing time. As such, only projects existing prior to the program - “HERA” and “HEROIC Santé”, got a score of at least 3/5.

JOGL awarded winners for each category a physical space for showcasing their project during the 2020 ChangeNow forum in the Grand Palais in Paris, and tickets for MaddyKeynote – a major innovation event in Paris. Two partners - Excelya and the Wild Code School, also provided rewards to the project of their choice. Additionally, the “Qualitative analysis of Tweets on Vaccination” was awarded to be the focus of a hackathon by the Wild Code School, and the project “APRICOT” was offered technical support for data science, legal and regulatory affairs by Excelya.

## Discussion

The Co-Immune programme was designed to foster the creation and collaborations of projects to address vaccination hesitancy and access to vaccination in France and around the globe. Despite this programme running for a short period of four months, we engaged a network of over 230 contributors who worked on 22 projects related to the programme challenges.

The use of on-site hackathons was beneficial in gathering non-academic participants from various backgrounds. Our data shows that in-person events and local outreach played a significant role in growing the community around Co-Immune. These offline events recruited 45% of the total community. Local enrolment was further strengthened by local partnerships, such as higher education organisations. While it was harder to contract partnership with public institutions in such a short time frame, the coordination team ensured alignment of the programme strategy with national and international policies by frequent consultation of public health bodies and mobilization of members of public institutions in its Committee for Ethics, Science and Impact. The development of new communities is usually a slow process in the absence of exogenous shocks such as the surge in collaborative communities created by the COVID-19 pandemic [52]. Tapping into existing projects and networks for events has proven to be fruitful in our case, allowing for a steady growth of the Co-Immune community.

While the offline hackathon-style events were important to build the community and create projects, the highly-rated projects which were eventually awarded did not originate or participate in those events, but rather benefited from Co-Immune as a platform for further growth. Several of these projects already existed before the start of Co-Immune and had a higher maturity level than the projects created during the short span of the programme. In addition, these projects were launched and run by people outside the larger Paris region. We thus stress the potential of online platforms and open innovation frameworks to build on existing projects, replicate, adapt and scale their activities in other contexts. This can be seen in the online events of Co-Immune, which were useful to reach worldwide participants. Recently, online events have been used widely during the COVID19 pandemic [52–54], supporting our initial assumption that forming and animating a distributed online community for public health programmes is a relevant approach.

Online platforms can also offer projects that started at hackathons a pathway to pursue their development after such events, potentially alleviating one of the main drawbacks of such short temporal interventions [42]. The sustainability of the newly created project efforts could potentially be improved using incentives such as microgrants or fellowship programmes for continuing projects in the post-programme period [55].

The use of JOGL platform enabled projects to gain visibility, list their needs to create interfaces for collaboration, and share open datasets, code and tools. In this case, it also enabled the programme coordinators to connect participants with project leaders based on a match between needs and skills. Yet, this approach was time-consuming, and scaling up our efforts proved to be challenging. The automation of such matchmaking tasks through a recommender system would help to minimize these efforts and increase the impact of projects through accelerated development [55].

At the governance level, the coordination team aimed at engaging beneficiaries and a wide range of collaborators as early as the design phase of the programme, to increase the relevance and sustainability of Co-immune’s potential outcomes. In our case, interviews of experts in immunization, literature review, and the contribution of health practitioners, helped develop the scientific orientation of the programme. This collaborator search led to the creation of an independent voluntary committee for Ethics, Science and Impact (CESI), which enabled projects to evolve within a clearly defined ethical framework. Mentoring is a known strategy that is used by open, online communities [56–58] and the Co-Immune programme made use of this strategy too. Given the diversity of backgrounds and level of expertise across the participants, it was necessary to engage a similar diversity within the mentors.

The benefits of our partnership framework were two-fold. Firstly, its agile nature enabled us to add partners as the community and its needs evolved. Through their expertise, these partners also enabled us to target prize categories that would best fit the type of projects emerging from the programme. Secondly, through their own experts and co-organizing events, several partners engaged in close relationships with JOGL and the individual projects. This was favorable to sustain collaborations and projects after the end of the programme.

## Conclusion

Co-Immune highlights how open innovation approaches and online platforms can help to gather and coordinate non-institutional communities in a rapid, distributed and global way towards solving SDG-related issues. The Co-Immune programme gathered participants and partners from various backgrounds in a newly formed community to facilitate the creation of new projects as well as the continuation of existing projects to address the issues of vaccination hesitancy and access. In an open framework, the projects made their data, code, and solutions publicly available.

Through the ideas of hackathons and other contest approaches, such initiatives can lead to the production and transfer of knowledge, creating novel solutions in the public health sector. The example of Co-Immune contributes to paving the way for organisations and individuals to collaboratively tackle future global challenges.

## Data Availability

Data is available in open access on Zenedo under the Licence: Creative Commons Attribution 4.0 International. 

https://doi.org/10.5281/zenodo.4560273

## Acknowledgements

First, we would like to thank all the Co-Immune participants that made the programme possible by bringing their creativity, skills and insights to address contemporary public health challenges.

We thank Sanofi for funding this programme and especially Diane Brément and Nansa Burlet for their assistance.

We thank the JOGL team for their work on coordinating the Co-Immune programme, with special efforts of Lola Casamitjana and Marine Vouard. We thank the Center for Research and Interdisciplinarity (CRI, Paris), Epitech Paris, Sup Biotech, and the Wild Code for their support in organizing events with their students; Kap Code, Excelya, Girls in Tech, CorrelAid, and Data for Good for their guidance and technical assistance to participants and projects; S3 Odeon, TUBA, Change Now, Maddy Keynote for the visibility they provided to this programme.

We thank the interviewees at the 7th Fondation Merieux Vaccine Acceptance conference for highlighting the key issues to address and potential solutions participants could build on. We thank the mentors for the support they provided to projects and participants throughout the duration of the Co-Immune programme. We thank Enric Senabre Hidalgo for insightful comments during the final stages of writing this manuscript.

Finally, we thank the members of the Committee for Science, Ethics, and Impact (CESI), Gilles Babinet, Jérôme Béranger, Anshu Bhardwaj, Liem Binh Luong Nguyen, Mélanie Heard, Ariel Lindner, Juliette Puret, Olivier Rozaire, for their valuable input that allowed the creation and implementation of a framework for ethics, science and impact for the Co-Immune programme and the independent assessment of its projects.

## Authors contributions

TEL and IV co-designed at the early stage of the initiative the thematic and the scope of the programme. CMM and TEL conceived the programme. CMM led the coordination team of the programme. CMM, BGT, TEL, CLBG and MS participated in the programme implementation. BGT, RJ, and MS analysed data and CMM, BGT, GF, CLBG, MS wrote the paper.

## Notes

### Competing Interest Statement

All authors have completed the ICMJE uniform disclosure form. No support from any organisation during the elaboration and submission work of this manuscript; Just One Giant Lab received funding from Sanofi  to develop and implement the programme which include publication fees. Sanofi respected the strict independence of JOGL, which administers its platform and the Co-Immune page in complete autonomy. Similarly, the Committee for Ethics, Science and Impact (CESI) was independent of Sanofi and decided alone on the strategic and scientific orientations of the program and the best projects to be rewarded.
GF was paid by Just One Giant Lab to support the elaboration of the manuscript; provided consulting services to Vaccines Europe, a trade association based in Belgium and is a volunteer board member of the Coalition for Life-Course Immunisation, a UK-based charity ; no other relationships or activities that could appear to have influenced the submitted work.

### Funding Statement

Just One Giant Lab received funding from Sanofi to develop and implement the programme which include publication fees.

### Author Declarations

Commission Nationale de l'Informatique et des Libertes registration number 2221728 (Methodologie de reference MR-004).

