## Supplementary information for "Co-Immune: a case study on open innovation for vaccination hesitancy and access"

#### Supplementary Method: comparing JOGL with other platforms

##### Selection of comparative platforms

In order to situate the action and suitability of JOGL as a host for Co-Immune, we selected popular social networks, science publishing and collaboration websites, and citizen science and project creation websites as elaborated on below.

*Social networks*: We selected platforms for which the primary focus is the connection of people and thoughts, without focus on projects or third person entities- they are dedicated to the self, and fulfil social needs, from chatting to self-promotion and networking (59): LinkedIn, Instagram, Quora, Facebook, Twitter, Discord, Reddit and Mind.

*Science publishing and collaboration site*s: These platforms are not focused on the self, but instead on third person goals such as manuscript edition and curation, project creation or grant acquisition.  We selected: Nature, Open Peer Review MNI, OSF collaboration, BioRxiv, pubpub, easychair, Je-s system, Research Gate, Academia, Mendeley and mySociety (RSB) (60–62).

*Citizen science and project creation sites*: These tools enable a commons based peer production, using open science, crowdsourced data collection, collaboration or challenge-based approaches: Kaggle, Hackaday, Experiment, Open Humans, Zooniverse, Instructables, Wikipedia, FlossManuals(63,64).

##### Social network feature comparisons method

We evaluated the presence (1) or absence (0) of 42 features across these platforms that were manually selected for their relevance with the organisation of the Co-Immune programme. Features encompass communication, collaboration, and participant behavior. Platform clustering was performed using correlation distance and average linkage method. In order to gain insights on the proximity between platforms and take into account correlation between features, we performed a principal component analysis (PCA) and projected platform feature vectors on the top two eigenvectors (Fig S1). This allowed us to visualise the 3 types of platforms (colors). The top eigenvector, concentrating most of the variance across features, segregates social networks from scientific and project-based platforms. The second component in turn separates publication from peer-production platforms. This allows us to situate JOGL in a central position with respect to the first component (mixing science and social network), while clustering it with peer-production platforms.

### Supplementary Table 1: Co-Immune partners

Supplementation Table 1: Co-Immune partners

| Partners | Status | Activity |
|---|---|---|
| Sanofi | Private | Healthcare Industry |
| S3 Odéon | Non-profit | Communication And Event |
| Epitech | Private | Education |
| CorrelAid | Non-profit | Innovation And Technology |
| SUP biotech | Private | Education |
| Excelya | Private | Research |
| Wild Code School | Private | Education |
| Girls in Tech | Non-profit | Innovation And Technology |
| Kap Code | Private | Innovation And Technology |
| TUBA | Non-profit | Incubator |
| Data for good | Non-profit | Incubator |
| Maddy Keynote | Private | Communication And Event |
| Change Now | Private | Communication And Event |

Supplementary Table 2: Project Assessment grid

### Co-Immune | Project Assessment

Congratulations for being part of our reviewers! You are going to play a very important role in the future of projects that could help humanity solve one of its great challenges, better health for all. Thank you!

Co-Immune is an open and collaborative research and innovation program carried out by the NGO Just One Giant Lab, which aims to contribute to improving vaccination coverage in France and across the globe. (More on app.jogl.io/program/coimmune)

Scores you attribute will be used by the honorary and independent Committee for Ethics, Science and Impact (CESI) of the Co-Immune program to attribute prizes on the 18th of December in Paris during the Co-Immune ceremony. Register for the event here http://bit.ly/co-immune_ceremony !

More information on the evaluation framework and prizes here: http://bit.ly/assessment-framework

WHEN?
This form is to be completed by the 16th or the 17th of December.

WHY?
This form will allow you to evaluate the projects submitted to the Co-Immune program, The scores and comments given will be used to assess the approach, implementation, and impact of each project.

WHAT?
We kindly request you to provide independent, fair, unbiased and constructive feedback. No conflict of interest with the Co-Immune financing partner shall be accepted.

HOW?
Assessment is made anonymously. JOGL will not share your name or contact information to any participant.
The comments given here will be made public on the platform. However, the given scores will be hidden to the public. Your identity will remain anonymous unless you purposely waive your anonymity.

QUESTIONS ?
If you have any questions, please, feel free to contact JOGL at

READY? GO!
* Required

1. Email address *

2. Your first name and last name *
This information is only used by JOGL for admin purpose and will not be made public. This evaluation is anonymous.

3. Please provide information regarding your current or previous links of interest with Sanofi, the main financial partner of the Co-Immune program. *

*Mark only one oval.*

- ◯ I certify I have no link of interest with Sanofi
- ◯ I have links of interest with Sanofi. Therefore, I regrettably cannot take part in the project assessment

4. Link of the project page being reviewed *
You can find the list of links and names of projects to review here: http://bit.ly/active-projects-links

**Project Approach**

In this section, you will assess the approach of the project. Please rank from 1 to 5 each criteria below. You can provide additional comments in the appropriate field.

5. The clarity and relevance of the problem that the project is attempting to solve and alignment with the Co-Immune program's scope [Score] *
1. Unclear and/or irrelevant | 3. Clear but relevant points are missing | 5. Very clear and relevant

*Mark only one oval.*

| 1 | 2 | 3 | 4 | 5 |
|---|---|---|---|---|
| ◯ | ◯ | ◯ | ◯ | ◯ |

6. The clarity and relevance of the problem that the project is attempting to solve and it's alignment with the Co-Immune program's scope [Comment, optional]

Please, comment and explain the score you have given. You can also give constructive feedback to help the project improve on this criteria.

7. Fit between the project's approach/methodology and the problem they have stated to resolve [Score] *

1. Inappropriate | 3. Appropriate | 5.Very appropriate

*Mark only one oval.*

| 1 | 2 | 3 | 4 | 5 |
|---|---|---|---|---|
| ⬭ | ⬭ | ⬭ | ⬭ | ⬭ |

8. Fit between the project's approach/methodology and the problem they have stated to resolve [Comment, optional]

Please, comment and explain the score you have given. You can also give constructive feedback to help the project improve on this criteria.

9. Introduction of ground-breaking objectives, novel concepts or approaches by the project [Score] *

1. Not innovative | 3. Innovative | 5. Disruptive

*Mark only one oval.*

| 1 | 2 | 3 | 4 | 5 |
|---|---|---|---|---|
| ⬭ | ⬭ | ⬭ | ⬭ | ⬭ |

10. Introduction of ground-breaking objectives, novel concepts and approaches [Comment, optional]

Please, comment and explain the score you have given. You can also give constructive feedback to help the project improve on this criteria.

#### Implementation

In this section, you will assess the implementation strategy of the project. Please rank from 1 to 5 each criteria below. You can provide additional comments in the appropriate field

11. The project's state of progress [Score] *

1. No results yet | 3. Some results but with a promising development plan | 5. Working proof of concept

*Mark only one oval.*

| 1 | 2 | 3 | 4 | 5 |
|---|---|---|---|---|
| ⬭ | ⬭ | ⬭ | ⬭ | ⬭ |

15. The project's ability to actively engage and align itself with all the relevant groups and possible stakeholders [Score] *

1. Unaware | 3. Aware but unclear integration | 5. Actively engaged

*Mark only one oval.*

| 1 | 2 | 3 | 4 | 5 |
|---|---|---|---|---|
| ⬭ | ⬭ | ⬭ | ⬭ | ⬭ |

16. Ability to actively engage and align itself with all the relevant groups and possible stakeholders [Comment, optional]

Please, comment and explain the score you have given. You can also give constructive feedback to help the project improve on this criteria.

_______________________________________________

_______________________________________________

_______________________________________________

_______________________________________________

_______________________________________________

#### Impact

In this section, you will assess the impact of the project. Please rank from 1 to 5 each criteria below. You can provide additional comments in the appropriate field.

17. Clarity and relevance of the criteria used to measure impact [Score] *

1. Unclear and irrelevant | 3. Clear but not entirely appropriate | 5. Very clear and relevant

*Mark only one oval.*

| 1 | 2 | 3 | 4 | 5 |
|---|---|---|---|---|
| ⬭ | ⬭ | ⬭ | ⬭ | ⬭ |

18. Clarity and relevance of the criteria used to measure impact [Comment, optional]

Please, comment and explain the score you have given. You can also give constructive feedback to help the project improve on this criteria.

_______________________________________________

_______________________________________________

_______________________________________________

_______________________________________________

_______________________________________________

19. To what extent does the project takes into account its ecosystem (ecological, environmental, ethical and social considerations) [Score] *

1. Unaware | 3. Aware | 5. Very aware

*Mark only one oval.*

| 1 | 2 | 3 | 4 | 5 |
|---|---|---|---|---|
| ⬭ | ⬭ | ⬭ | ⬭ | ⬭ |

20. To what extent does the project takes into account its ecosystem (ecological, environmental, ethical and social considerations) [Comment, optional]

Please, comment and explain the score you have given. You can also give constructive feedback to help the project improve on this criteria.

_______________________________________________

_______________________________________________

_______________________________________________

_______________________________________________

_______________________________________________

21. The sustainability and scalability of the project in the long term [Score] *

1. No sustainability model and no scaling potential | 3. Sustainable and scalable but still unstructured plan | 5. Already applying a sustainable plan & good scalability

*Mark only one oval.*

| 1 | 2 | 3 | 4 | 5 |
| --- | --- | --- | --- | --- |
| ⬭ | ⬭ | ⬭ | ⬭ | ⬭ |

22. The sustainability and scalability of the project in the long term [Comment, optional]

Please, comment and explain the score you have given. You can also give constructive feedback to help the project improve on this criteria.

23. The project's dissemination strategy (quality of documentation for goals, results, methods and needs; open access model, outreach) [Score] *

1. Irreproducible and poorly documented 3. Good documentation but with few weaknesses 5. Easily reproducible & very well communicated

*Mark only one oval.*

| 1 | 2 | 3 | 4 | 5 |
| --- | --- | --- | --- | --- |
| ⬭ | ⬭ | ⬭ | ⬭ | ⬭ |

24. The projects dissemination strategy (quality of documentation for goals, results, methods and needs; open access model, outreach) [Comment, optional]

Additional comments and reviewer self-evaluation

You may provide below additional comments addressed to the team leaders of the project, and private comments to the JOGL team.

Please, also assess your expertise in the fields relevant to evaluate the project.

25. [To the team leaders] Other general constructive comments about the project *

26. [To JOGL] Private comment - anything you would like to inform the JOGL team of, privately.

27. What is your expertise that is relevant to evaluate this project? *

Please provide justification of your expertise if possible.

28. Would you like us to include you in our "Experts community" and provide similar reviews to projects when you have time in the future? *

*Mark only one oval.*

- Yes
- No
- Maybe

Thank you very much for your feedback!

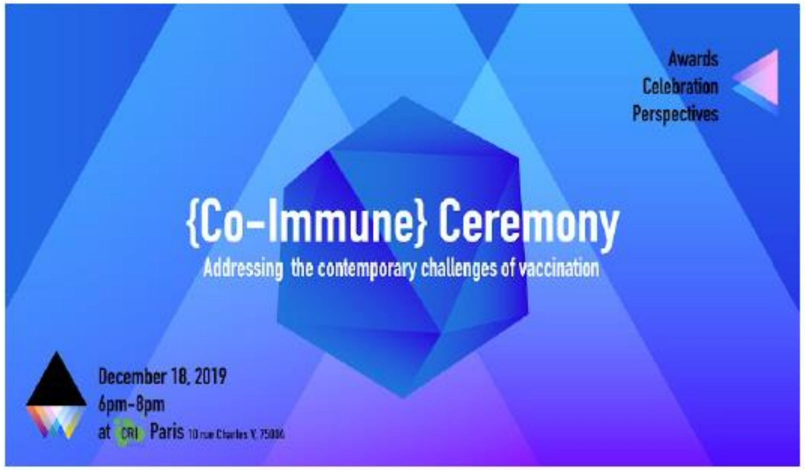

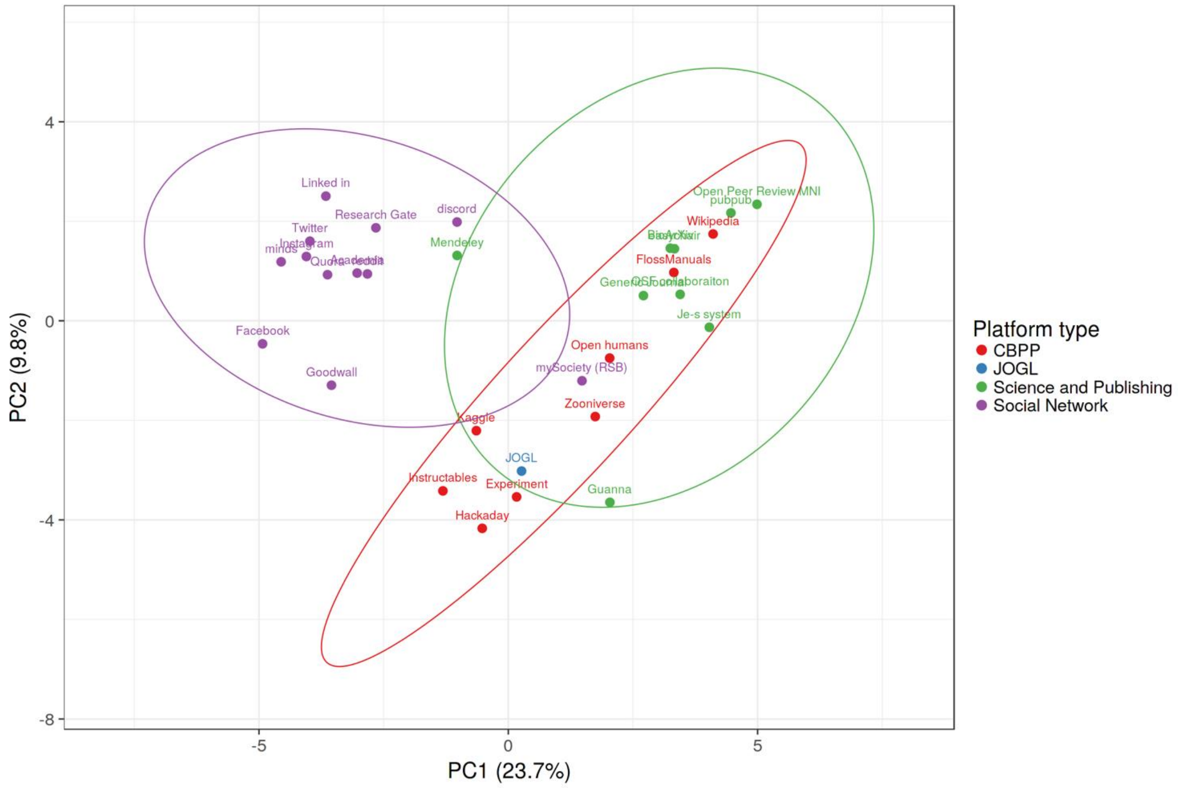


**Supplementary Fig 1: JOGLs position as a social network for Open Science.**

Various online platforms represented in the first 2 components of a PCA based on their features from Fig1. We use the pcaMethods R package with default parameters to calculate principal components. Catchment of groupings represent a 95% certainty interval of a platform landing within the platform type through feature space. N = 29 platforms, each with 42 true/false data points. CBPP- Citizen based peer production network
